# Dengue Forecasting Models: A Systematic Review and Network Meta-Analysis of Relative Performance

**DOI:** 10.64898/2026.02.18.26346534

**Authors:** Pitak Benjarattanaporn, Debebe Adewo, Anthea Sutton, Andrew Lee, Peter J. Dodd

## Abstract

**Background:** Accurate dengue forecasting is vital for public health preparedness. Despite a surge in forecasting approaches, a quantitative ranking of the relative performance and practical utility of dengue forecasting is lacking.

**Methods:** A systematic review and Network Meta-Analysis (NMA) of studies comparing dengue forecasting methods (2014–2024) was conducted. Models were categorised into five groups: Time Series, Deep Learning (DL), Machine Learning (excluding DL), Hybrid, and Ensembles. NMA was applied to the logarithm of the most common forecast error metric to rank relative performance—an “Implementability Score” quantified analyst and data requirements, and computational costs.

**Results:** 59 studies were included. NMA of Root Mean Squared Error identified k-Nearest Neighbour (k-NN) models as achieving the highest predictive accuracy, followed closely by Vector Autoregression, Kalman Filtering, Generalised Linear Model and Autoregressive Neural Network (ARNN). While DL models showed high potential, they scored lowest in implementability due to poor interpretability and high data requirements. Most studies utilised meteorological covariates, with significant gaps in the use of socio-economic and entomological predictors.

**Conclusions:** Although there was some trade-off between accuracy and implementability, traditional statistical models were often comparable in accuracy to machine learning approaches, with advantages in interpretability and data needs. Under-explored areas for future research include the use of ensemble models and the use of socio-economic and entomological data.

**Registration:** PROSPERO CRD420251016662.

**Author Summary:** Dengue is a critical global health threat affecting the world’s population. While many forecasting models exist to help officials prepare for outbreaks, there has been no standardised way to compare their performance. This leaves health experts in resource-limited areas uncertain about which tools are truly reliable or easy to use under their specific local conditions.

We conducted a network meta-analysis of studies comparing dengue forecasting methods’ accuracy, grouping them into five categories: Machine Learning, Deep Learning, Time Series, Ensemble, and Hybrid. Beyond ranking their accuracy, we developed an “Implementability Score” to evaluate the practical feasibility of each model, accounting for technical complexity, data requirements, and software accessibility.

Our analysis identified the top-performing models. Notably, traditional statistical models often performed as well as complex Deep Learning algorithms. While advanced models show potential, they are often difficult to implement or explain to decision-makers.

There is no “one-size-fits-all” solution; the best model depends on capacity and data in each setting. This study provides a roadmap for public health officials to select tools that are both accurate and feasible.

## Introduction

Dengue is a major global health challenge with approximately half the world’s population at risk across over 100 endemic countries [1]. The burden of dengue is increasing both in terms of case numbers and socioeconomic impact. Dengue is a mosquito-borne viral disease caused by four serotypes and is transmitted by the *Aedes aegypti* mosquito. Annually, an estimated 390 million dengue infections occur globally, with 96 million manifesting clinically [2]. The disease remains endemic predominantly in tropical and subtropical regions, where climate change, urbanisation, and insufficient mosquito control contribute to its persistent transmission. This high disease incidence places a strain on healthcare systems, leads to loss of productivity, and diminishes the quality of life for affected populations. Consequently, strategies emphasising disease surveillance, such as effective dengue case monitoring systems, are essential tools for accurately tracking and controlling the disease locally [3]. While Dengue often presents as a mild, self-limiting febrile illness, dengue can progress to severe forms, including dengue haemorrhagic fever that can lead to plasma leakage, severe bleeding, or multi-organ impairment, posing a particular threat to older adults and those with a secondary dengue infection.

The increasing volume and sophistication of data collection methods within modern surveillance systems have enabled the recent surge in dengue forecasting models [4]. As advances in information technology and epidemiological modelling have improved the quality and accessibility of these datasets, there has been a growing emphasis on translating this data into accurate, actionable insights for public health response [5,6]. Given the cyclical nature of dengue epidemics, forecasting plays an important role in mitigating the public health impact of the disease. The ability to predict dengue outbreaks and estimate the magnitude and timing of impending case surges is important for proactive public health response planning [7]. Forecasting enables public health sectors to anticipate and manage the demand on healthcare systems [8]. Specifically, these predictions enable authorities to allocate resources, such as the number of healthcare professionals and medical supplies (e.g., testing kits, vaccines), in advance of the peak transmission period [9,10]. Forecasting models provide the necessary lead time for the optimal timing of interventions. Predictive alerts can trigger targeted, control measures such as insecticide fogging or community campaigns aimed at eliminating mosquito breeding grounds.

In this study, we focus on disease forecasting as the prospective process of estimating the future incidence and outbreak. Importantly, we exclude other meanings of ‘prediction,’ a term often applied to individual-level clinical outcomes or to the estimation of contemporary disease levels in areas where observational data is missing. In such spatial contexts, prediction utilises statistical models to infer current prevalence across a geography based on available environmental or demographic data. Forecasting, in contrast, takes a population-level probability approach, assigning a probability to a particular future trend [8,11].

A wide range of methods has been employed in forecasting models for dengue, from basic statistics to advanced machine learning. However, it is unclear which models work best for a specific area or context. Existing reviews of dengue prediction modelling have focused less on forecasting and have not made quantitative comparisons between models. Previous systematic reviews have addressed distinct dimensions, including the integration of social media and machine learning [12], the analysis of climatic predictors [13], and the identification of optimal machine learning models [14]. Our aim in this study is to guide analysts on what methods exist to forecast dengue, and what their relative merits are. We therefore systematically identified studies that compared methods’ performance on dengue time series, and applied Network Meta-Analysis (NMA) to quantitatively compare and rank their performance. Recognising the key importance of practical utility, we also conducted an implementability assessment of the top-performing models to generate practical recommendations for analysts and modellers.

## Methods

We followed the Preferred Reporting Items for Systematic Reviews and Meta-Analyses for network meta-analyses (PRISMA-NMA) 2015 guidelines for conducting and reporting the systematic review [15]. A systematic review protocol was developed and registered in PROSPERO (<u>CRD420251016662</u>).

### Study selection process

Inclusion criteria focus on studies that forecast dengue incidence and compare the performance of two or more forecasting methods using quantitative metrics. All forms of dengue data representing population-level incidence were admissible, including clinically suspected and laboratory-confirmed cases. Studies that develop or apply a single forecasting method without comparing it to others were excluded. Studies that included predictions for infectious diseases other than dengue were excluded. Furthermore, studies focusing on prediction rather than forecasting were also excluded. Accordingly, we included studies that quantitatively evaluated the relative performance of multiple forecasting methodologies to dengue incidence data. Excluded from the review were studies that did not involve comparing forecasting methods on time series data.

Only published, peer-reviewed articles and conference papers in the English language were included in this review. Grey literature, books, chapters, dissertations, and preprints were excluded. Our review question and subsequent eligibility criteria were developed using a modified PICO framework. The standard PICO model, commonly used for clinical intervention reviews, was adapted to align with the unique characteristics of forecasting methods. Our modified framework is defined as PICOS to better suit the nature of our research question, which focuses on comparing diverse forecasting methodologies [16]. The search strategy and inclusion/exclusion criteria were defined using the PICOS framework. A detailed summary of the PICOS criteria is provided in Appendix Table A1.

The search results were exported into Zotero for reference management, where duplicates were identified and eliminated. Following deduplication, the references were imported into the Rayyan platform [17], where further duplicates were removed before screening. Two reviewers assessed the Titles and abstracts (PB and PD), while three reviewers evaluated the full texts (PB, PD, and DS). PB resolved any conflicts that arose with consensus.

### Search Strategy and Literature Search

A literature search was conducted on 7 March 2025 using the electronic databases MEDLINE, Scopus, and Web of Science to obtain articles on forecasting the incidence of dengue. The search was for articles published from 1 January 2014 to 7 March 2025. We restricted the literature search to journal articles and conference papers published in English.

The search strategy focused on three core themes: Dengue, Forecasting, and statistical modelling. The search was also limited to a specific article publication year range (Appendix B1).

### Data extraction & Narrative summary

A data extraction tool based on Google Spreadsheets was utilised for the studies included in the analysis. The extracted data encompassed various aspects, including study publication type and temporal distribution, the programs or programming languages employed, covariates used, data sources, granularity of time series data, forecasting models, model validations, forecast time horizon, and reported performance metrics utilised, which were summarised in this review.

For the narrative synthesis, we grouped studies based on their methodological approach. We identified and categorised the forecasting models according to their unique features, such as Time Series (TS), Deep Learning (DL), Machine Learning (ML) exclusive of deep learning architectures, Hybrid, and Ensemble models. The findings were shared in a narrative style, with the extracted data displayed through tables.

We distinguished between hybrid and ensemble methodologies. Hybrid models were defined as integrating heterogeneous techniques into a single, interdependent framework [18]. Ensemble models were defined as the aggregation of multiple independent model outputs using statistical combiners [19].

### Quality assessment

The quality of the included studies was assessed using an adapted version of the Epidemic Forecasting and Generalizability (EPIFORGE) checklist [20,21]. This evaluation focused on key methodological domains for the validity and interpretability of epidemic forecasting research, examining the reporting of study goals, data sources, model characteristics and underlying assumptions, the rigour of model evaluation procedures, and considerations of study generalizability. Nine equally important criteria were scored on a three-point scale ranging from 0 (poor reporting), 1 (Fair reporting) to 2 (good reporting), yielding a maximum achievable quality score of 18. Based on the cumulative score, each included study was subsequently classified into one of four quality categories: ‘low’ (< 10), ‘medium’ (10–12), ‘high’ (13–15), or ‘very high’ (> 15), providing a structured framework for understanding the methodological strength of the evidence synthesised in this review. The quality assessment was performed by PB and reviewed by PD and DS.

### Quantitative Synthesis of Model Performance

The primary goal of the data synthesis was to compare the predictive performance of the models.

We extracted all metrics of model forecast error from each included study and identified the most commonly reported metric of forecast errors. This metric was designated as the primary outcome variable for the Network Meta-Analysis (NMA). NMA extends traditional meta-analysis by working with an evidence network of comparisons between multiple options to estimate their relative performance from both direct and indirect empirical comparisons [22].

In order to ignore the specific magnitude or scale associated with individual datasets, we applied a natural logarithm transformation to predictive error metrics. We restricted ourselves to studies using the most common predictive error metric. Our meta-analysis thus estimated the relative accuracy between any two models in the network [33].

An NMA network diagram was constructed using the extracted data to visually represent the direct and indirect comparisons available between the different forecasting model categories. We reported NMA results relative to the ‘Naive’ model as the reference baseline. This model was defined using a Last Observation Carried Forward (LOCF) approach [23], where the forecast for a given period is simply the value observed in the most recent preceding period. The Naive model was chosen in the expectation that more complex modelling approaches would outperform it.

The NMA was fitted in a Bayesian paradigm using the multinma package (version 0.8.0) in R software (version 4.4.1). [24,25]. Specifically, we applied weakly informative normal priors for all parameters and ran the MCMC simulations for 4,000 iterations. Convergence was verified using R-hat statistics and trace plots, while the Deviance Information Criterion (DIC) was used to evaluate the overall model fit. For each method, we output indicators of relative performance, including the relative effect [26], the predictive ranking, and the Surface under the Cumulative Ranking (SUCRA) values [27].

### Implementability Assessment

We assessed the model characteristics for the top five highest-performing models identified in the Quantitative Synthesis across two key dimensions: ease of implementation and resource requirements. The assessment of ease of usability encompassed assessing 1) Availability of open-source Python/R packages, 2) Transparency/Interpretability, and 3) Complexity of parameter tuning. Resource requirements involved assessing the resources necessary for model deployment, focusing specifically on 4) Computational Requirements and 5) Input Data Requirements.

This qualitative assessment used a four-point ordinal scale to classify model requirements and ease of use: Very High, High, Moderate, and Low. Detailed operational definitions are provided in Appendix D, Table D1. For most dimensions (i.e. Data Requirements, Computational Requirements, and Complexity), a low score is the most desirable outcome, indicating the lowest effort or resource demand for parameter tuning, data needs, or computation, whereas a high score is undesirable, signifying the highest complexity, demand, or resource intensity. However, for the Model Availability (open-source Python/R packages) and Interpretability dimensions, high scores are positive, meaning the model is readily available and maintained, or that it provides a clear relationship between covariates and predictions. Conversely, low scores are negative, requiring full custom code reproduction or offering no practical way to explain the underlying logic. The scale is visually represented using a red-to-green gradient, where green represents the most feasible outcome and red represents the least feasible outcome.

## Results

The initial database search generated 4,791 records; the final review included 59 papers. Figure 1 presents the PRISMA flow diagram. This section summarises the findings of the review. The full list of papers and their properties is provided in Appendix B, Table B1.

**Figure 1.**
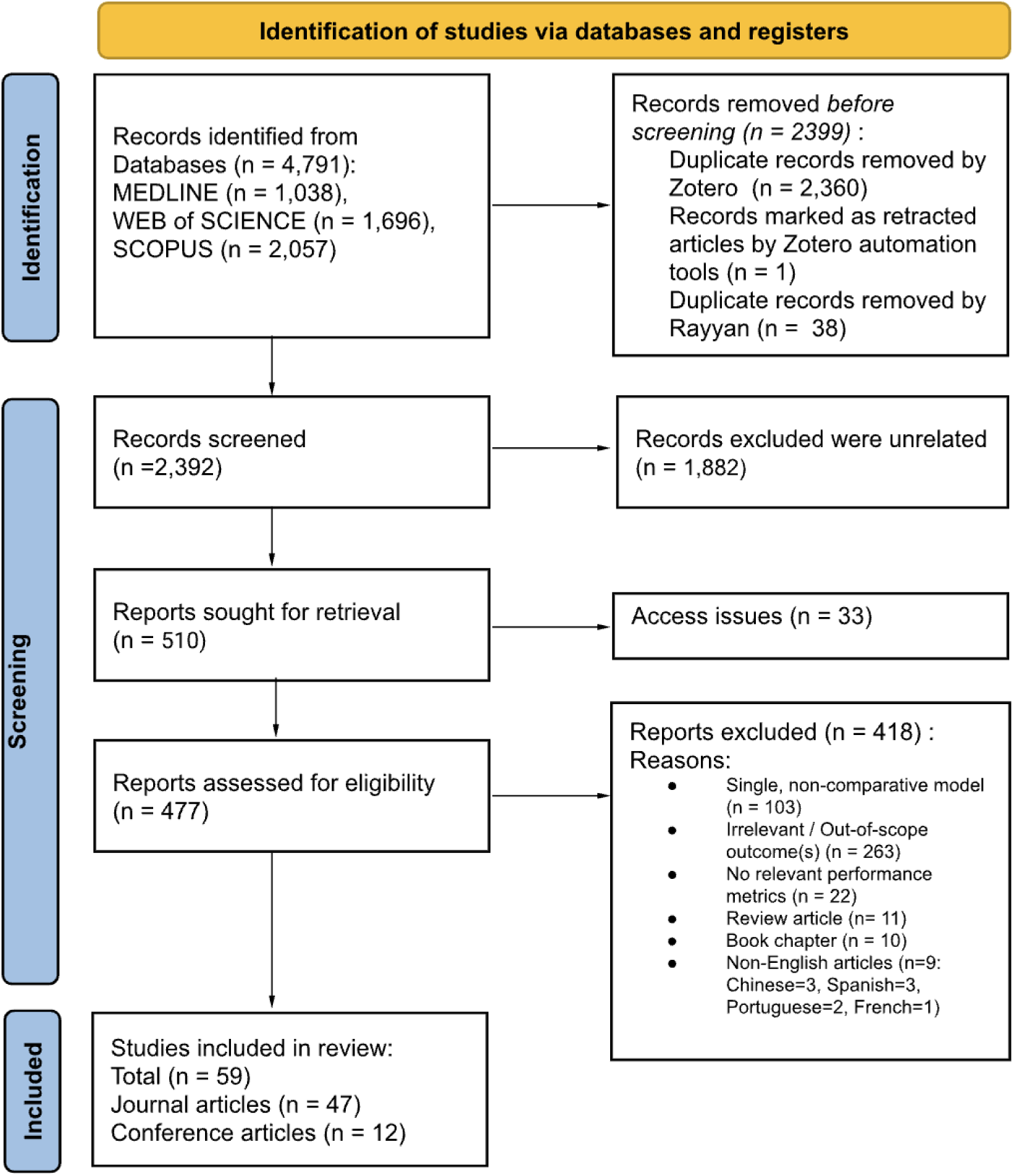
PRISMA Flow Diagram Depicting the Study Selection Process.

### Distribution of Dengue Forecasting Articles

The temporal distribution of the 59 articles included in this review covers the period from 2014 to 2025. The number of journal articles shows a consistent upward trend throughout the observed period, peaking in 2020 with a total of 10 publications. This review encompasses studies conducted across 20 distinct countries, with a significant concentration in Asia, followed by Latin America.

### Data Types

All included studies (100%) incorporated epidemiological covariates, followed by meteorological covariates (n=46, 77.97%). Geospatial, demography (both n=10, 16.95%), and internet data (n=7, 11.86%) were also used. The least common were entomology and socio-economic factors (both n=2, 3.39%). The primary predictor was typically the case count. The majority of studies (92.19%) relied on notified incidence data, primarily representing clinically suspected or laboratory-confirmed cases extracted from governmental surveillance registries. In terms of temporal resolution, half of the included studies utilised data aggregated at the weekly level.

### Forecasting model

Forecasting methods were categorised into five main groups based on the number of included studies that implemented at least one model from that category. The distribution was: Machine Learning (ML) (n=48 studies, 38.1%), Time Series (TS) (n=36 studies, 28.6%), Deep Learning (DL) (n=23 studies, 18.3%), Hybrid (n=11 studies, 8.7%), and Ensemble (n=8 studies, 6.3%). The three most frequently implemented specific models, based on the count of studies, were Linear Regression (LR) (n=21 studies) from the ML group, ARIMA (n=20 studies) from TS, and Long Short-Term Memory (LSTM) (n=19 studies) from DL.(see Appendix Table 2B for the full data set)

The analysis of model comparisons is summarised in the network diagram (Figure 2). The size of each node is scaled according to the total number of times a model type is included in a comparative analysis. Machine Learning is the most common category, forming the largest node with 918 entries, followed by Time Series (449 entries) and Deep Learning (404 entries). The edge weight represents the number of direct head-to-head comparisons. The network indicates that the most frequent comparisons occurred equally between Time Series (TS) and Machine Learning (ML) models (n=27 comparisons) and ML and Deep Learning (DL) models (n=27 comparisons), with comparisons between TS and DL models (n=21 comparisons) being the next most frequent.

**Figure 2.**
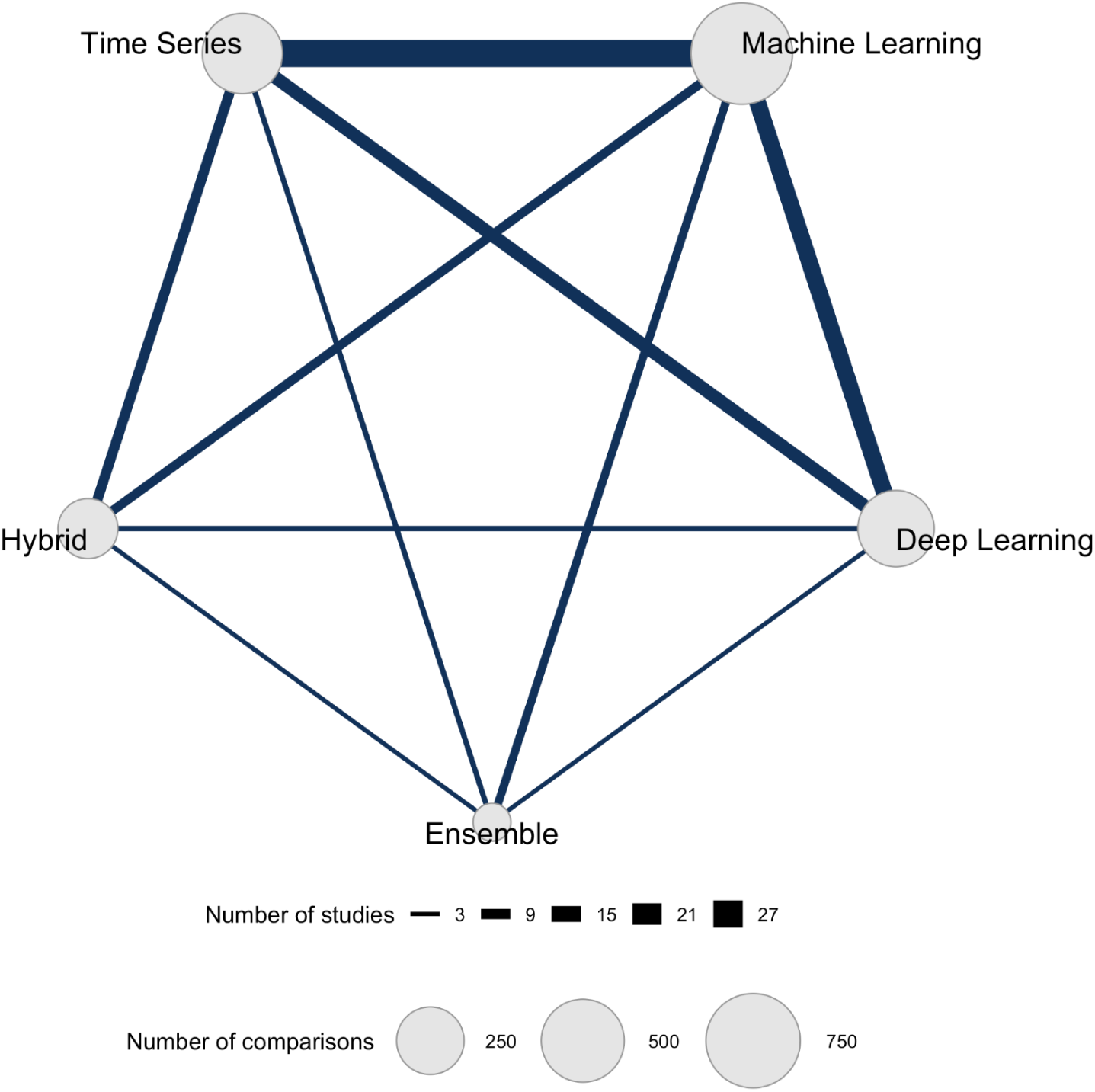
Network analysis of the forecasting model categories.

### Data Type by Model Type

Figure 3 illustrates the quantitative relationship between the covariate categories utilised and the forecasting model types employed across the included studies. The heatmap’s count represents the total number of times a specific Model Type and Covariate Category combination was utilised across all studies. Within the forecasting space, Machine Learning algorithms represent the dominant utilisation. The single most frequent combination observed is the integration of Meteorological data with Machine Learning (2,727 usages), followed by the utilisation of Meteorological data with Deep Learning (1,164 usages).

**Figure 3.**
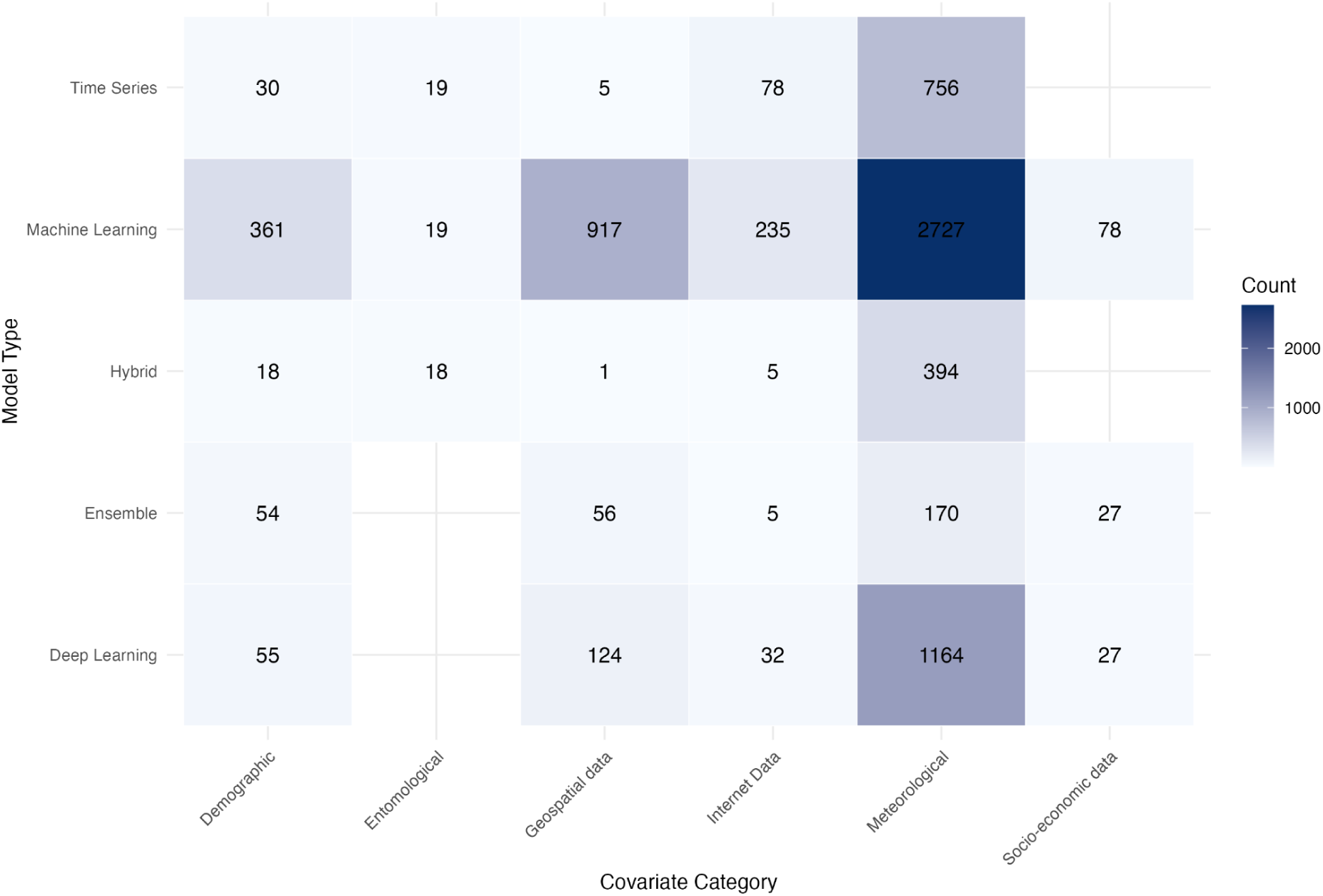
Heatmap of frequency of usage of specific Covariate Category and Model Type combinations across all studies. The count represents the total number of times each specific pairing of model architecture and data input was implemented within the included studies.

### Quality assessment

The adapted tool for assessing the reporting quality of forecasting studies yielded a score range from 6 (low quality) to 18 (very high quality) for the included papers. Of the studies evaluated, 12 were classified as low quality, 17 as medium quality, 22 as high quality, and 8 as very high quality. The average score was 12.3 out of 18, categorising the overall quality as high. All studies clearly defined the country of study. Discussions about data limitations were absent in 35 of the 59 analysed studies. (Appendix D, Table D3).

### Quantitative Synthesis

The three most frequently reported model performance evaluation metrics in dengue incidence forecasting studies were Root Mean Squared Error (RMSE) (n=47, 79.66%), Mean Absolute Error (MAE) (n=37, 62.71%) and Mean Absolute Percentage Error (MAPE) (n=12, 20.34%) (appendix C, Table C2). While RMSE, MAE, and MAPE were the most common, some studies used other metrics or combinations. Studies (n=6) [28–33] reported MAE but not RMSE. Additionally, many studies (n=5) [34–38] utilised highly specific evaluation metrics. Most of the selected studies commonly utilised Root Mean Squared Error (RMSE) as an evaluation metric.

The NMA model demonstrated an adequate fit to the data, with a residual deviance of 1393.9 compared to 1,556 data points. The effective number of parameters (pD) was 850.2, resulting in an overall Deviance Information Criterion (DIC) of 2244.1.

Figure 4 shows the NMA relative effect for each model compared to the Naive model baseline (indicated by the vertical red dashed line at 0). Models with a negative relative effect are statistically superior to the baseline. The Naive model represents the poorest performing method in our analysis.

**Figure 4.**
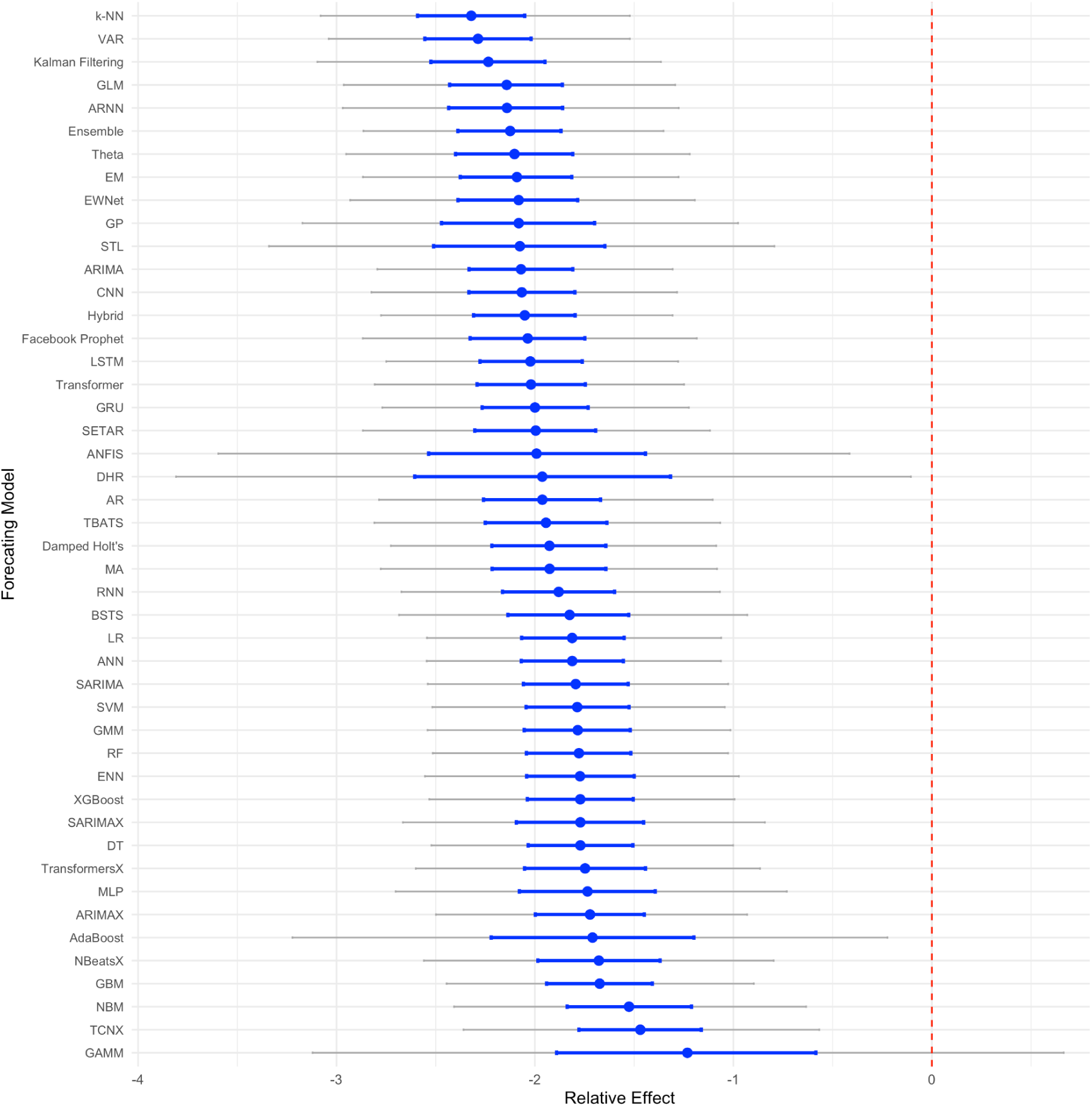
Comparative forecasting accuracy from network meta-analysis. Smaller relative effects indicate better fits. Effects are measured with respect to the “Naive” model.

Many methods showed comparable performance. The analysis identifies the k-Nearest Neighbours (k-NN) model as demonstrating the highest point estimate for relative accuracy, with a mean relative effect of -2.32 (95% CrI: -3.08 to -1.52). This was followed by Vector Autoregression (VAR) (RE: -2.29; 95% CrI: -3.04 to -1.52) and Kalman Filtering (RE: -2.24; 95% CrI: -3.10 to -1.37). Other high-performing methodologies included Generalised Linear Models (GLM) (RE: -2.14; 95% CrI: -2.96 to -1.29), Autoregressive Neural Networks (ARNN) (RE: -2.14; 95% CrI: -2.97 to -1.28), and Ensemble methods (RE: -2.13; 95% CrI: -2.86 to -1.35).

To determine the hierarchy of model performance, we calculated the SUCRA for each methodology. The k-NN model was identified as the top-ranked forecasting approach with a SUCRA of 90.26%, followed by VAR at 88.44%. Kalman Filtering at 79.08% and Ensemble models at 75.88% followed in the rankings. Despite the competitive mean relative effects observed for ARNN and GLM, their SUCRA scores were at 74.13% and 72.94%, respectively. The Additional details can be found in Appendix C, Table C1, and the rankogram for all included interventions is presented in Figure C1.

To synthesise the Implementation Assessment, the top five performing forecasting models identified from the Network Meta-Analysis (NMA) relative performance—specifically k-NN, VAR, Kalman Filtering, GLM, and ARNN were evaluated through a systematic implementability assessment (Table 1). The results are summarised in Table 1 (details of the qualitative scoring are provided in Appendix D, Table D1). The analysis showed a consistent trend where high predictive accuracy is often associated with medium-to-low overall feasibility. For example, the models with the highest mean performance are k-NN (Rank 1) and VAR (Rank 2). This classification is primarily driven by k-NN’s High score for Data Requirements and VAR’s Moderate requirement for Complexity of parameter tuning. ARNN (Rank 5 in performance) was the least feasible of the top five, requiring a Very High score for the Complexity of parameter tuning and Data Requirements. In contrast, models such as the GLM and Kalman Filtering were rated as highly feasible, benefiting from Low scores across both computational requirements and parameter tuning complexity.

**Table 1.** Performance and Implementability of Top-Performing Models.

| Rank | Forecasting Model | Ease of Implementation |  |  | Resource Requirement |  |
| --- | --- | --- | --- | --- | --- | --- |
|  |  | Availability of open-source Python/R packages | Transparency/Interpretability | Complexity of parameter tuning | Computational Requirements | Data Requirements |
| 1 | k-NN (k-Nearest Neighbours) | Very High | Moderate | Moderate | Low | High |
| 2 | VAR (Vector Autoregression) | Very High | Very High | Moderate | Low | Moderate |
| 3 | Kalman Filtering | Very High | High | Low | Low | Low |
| 4 | GLM (Generalised Linear Model) | Very High | Very High | Low | Low | Moderate |
| 5 | ARNN (Autoregressive Neural Network) | High | Low | Very High | Moderate | Very High |

### Reported Programming Language and Software

Among the 59 studies analysed, 12 (20.23%) utilised R software [33,39–49], while 7 (11.86%) employed Python software [31,42,50–54]. Additionally, only one study (1.69%) utilised both SPSS and MATLAB [55]. The remaining 39 (66.1%) of studies did not specify the software used.

## Discussion

This review systematically explored dengue incidence forecasting studies, focusing on studies comparing forecasting methods for time series published since 2014. By using NMA, we were able to meta-analyse the relative forecast accuracy of methods used, finding that k-NN, VAR, Kalman Filtering, GLM and ARNN had the best predictive accuracy. Our implementability assessment suggests that while software packages are easily available to implement these 5 best-performing methods, k-NN and ARNN require more data and provide less transparent and interpretable results.

Our findings extend and refine the conclusions of previous systematic reviews of dengue prediction methods. To navigate the current literature, researchers seeking long-term forecasting regarding climate change scenarios should consult Islan et al (2025) [56], while those interested in the integration of social media and internet search queries should refer to Sylvestre et al. (2022) [12]. For the use of satellite imagery in forecasting, the framework of the dengue incidence forecasting pipeline and the categorisation of forecasting models, Li and Dong (2022) [57], is a key resource. Our findings align with Leung et al. (2023) [13] and Fang et al. (2024) [14] regarding the growing use of machine learning and the central role of weather data. However, our study offers a different perspective on quality assessment. While Lim et al. (2023) [21] utilised the ‘EPIFORGE’ framework to evaluate methodological quality, we identified a need for a bespoke, adapted EPIFORGE specifically designed to capture the unique biases inherent in time-series forecasting. Additionally, we go beyond these assessments to include an ‘implementability assessment’, which can inform thinking around models’ technical complexity and the actual resources available to health practitioners.

Comparing our meta-analytic ranking of predictive performance (Figure 4) with the implementation assessment (Table 1) shows that there are many highly feasible methods (GLM, VAR, and Kalman Filtering) that are expected to perform well and are comparable to the best approaches. However, two of the top five ranked approaches used machine learning methods, k-NN and ARNN, both of which required larger volumes of training data and yielded models that lacked transparent interpretations. The majority of publications on these methods have been within the last 5 years. Given the advancements in machine learning methods and the increasing volume of data, this trade-off between interpretability and predictive performance is likely to become a growing consideration. Work to help understand and interpret machine learning models is a developing area, and may help ameliorate this issue [58]. In applied use, the relative value placed on accuracy versus transparency is a question for the commissioners of such modelling, and stakeholder engagement work should be considered to elicit preferences when investing in developing forecasting pipelines. However, in many practical cases, the data available for model development may render performance gains from machine learning approaches small compared to well-performing traditional approaches with higher interpretability. Ultimately, the choice of model should be informed by the intended public health use rather than statistical performance alone. For strategic resource allocation, higher complexity models with longer lead times are appropriate. However, for tactical local interventions, simpler models that can be run on basic hardware with immediate data entry are likely to be more feasible [59].

Notably, among the top-performing models, the GLM and ensemble methods ranked differently using relative accuracy than using SUCRA values. While GLM achieved a slightly more favourable mean relative effect value, ensemble models demonstrated higher SUCRA values. This statistical nuance suggests that although GLMs can forecast effectively in specific contexts, they may also exhibit greater variance across different datasets. In contrast, the higher SUCRA for ensemble models indicates a more consistent performance ‘near the top’ across the varied conditions included in this review.

Furthermore, we found that Hybrid and Ensemble models were much less explored areas compared to Time Series, Machine Learning, and Deep Learning models. Given their relatively strong performance, especially when ranked by SUCRA, and anticipated strengths in combining the strengths of different approaches, future research should include these methodological combinations in comparative assessments of forecasting.

The review found that data availability directly dictates model selection, as the structural characteristics of available data often determine the mathematical framework required. Dengue forecasting heavily relies on conventional epidemiological and environmental variables; the underutilisation of socio-economic and entomological variables represents a gap in the literature [34,35,38,44,60,60–64]. This finding is consistent with other reviews on the different aspects of dengue forecasting. Li and Dong highlighted the lack of studies that integrate socio-economic data into the predictive frameworks [57]. Incorporating these less common data sources, particularly in Hybrid or Ensemble frameworks, represents a promising direction for improving predictive capacity by capturing the complex, multi-faceted nature of dengue transmission that extends beyond purely climatic factors [65]. Dengue incidence case surveillance data were most commonly available at a weekly scale, followed by monthly and daily granularities [14]. It is important to note that a higher granularity of data is generally more beneficial for the timely detection of outbreaks.

While our overall assessment of the included papers had a generally high quality in reporting, roughly half of the included papers failed to specify the source of their data, hindering replicability and transparency. This lack of transparency extends to the computational tools used; a significant majority of studies failed to report the software or programming language employed in their analysis, among the minority that did disclose this information. This issue is particularly acute for more complex methodologies, such as Hybrid and Ensemble models, which often require custom scripts to integrate disparate algorithms.

This review has four main limitations that should be considered when interpreting our findings:

First, combining findings from different forecasting studies is challenging. The quality and availability of data vary significantly. For example, surveillance data differ in detail and completeness across regions, environmental data comes from various sources with different resolutions, and socio-demographic data can be outdated [34,60]. Furthermore, the field uses many different modelling approaches. Each of these models has its assumptions and limitations, which makes direct comparisons difficult [66].

Second, standardised tools for assessing the quality and risk of bias in forecasting research are currently lacking [6]. We needed to develop our framework to evaluate the quality of the studies included in this review [21]. This highlights a critical need for the development of better, standardised tools to assess the methods and potential biases in future dengue forecasting studies.

Third, our quantitative synthesis of model performance is restricted only to studies reporting the RMSE metric. Studies that reported only alternative performance metrics could not be included in the comparative analysis, which may lead to an incomplete representation of the full range of model performance.

Finally, the Performance and Implementability of the assessed models were inferred from the literature rather than through direct consultation with end-users. This study did not survey public health practitioners regarding their specific experiences with ease of development and use.

The principal limitation of our meta-analytic approach to ranking methods is the potential confounding of method with data and also with analyst skill. We noted that some methods tend to be used in conjunction with particular types of additional data. In these cases, it is impossible to completely separate performance gains due to methodology from those due to additional data. Where skill or experience is involved in the effective use of certain methods, it is not possible to separate differences in performance that are due to analysts rather than methods. Indeed, more skilled analysts may be drawn to choose certain methods, which could be both intrinsically better in terms of forecasting accuracy and more difficult to use well.

Our study demonstrated the potential for NMA to generate rankings of multiple options from incomplete and repeated sets of comparisons, outside of its usual area of application (i.e. analyses of clinical drug trials). NMA could be used to compare the predictive performance of forecasting methods for other infectious diseases where no complete set of comparisons has been made. However, the feasibility of such cross-study synthesis is currently hampered by inconsistent reporting.

The field would benefit from standardised reporting protocols, specifically regarding error metrics, clear definitions of forecasting horizons, and the mandatory description and sharing of hyperparameters and code, thus allowing for more meaningful performance comparisons across different outbreak scales and geographical regions [67]. In addition to this, the development of open-source, standardised datasets where different algorithms could be tested using the same data and metrics would allow clearer comparisons of performance and computational expense. Finally, we recommend that future research employ qualitative methods to establish how commissioners and analysts of applied work value accuracy and the human and computational resources required to develop forecasting pipelines.

## Conclusion

Our analysis identified a set of dengue forecasting methods with high relative accuracy, which exhibited some trade-offs between accuracy and implementability. Although the use of machine learning is expanding, traditional statistical models are often comparable in accuracy, generate interpretable results, and may require less data. Under-explored areas for future research include the use of ensemble models, and the use of socio-economic and entomological data.

## Supporting information

Supplementary Material 2: PRISMA Checklist

## Data Availability

All data extracted and analysed during this systematic review are included in the manuscript and its supplementary information files. This study is a synthesis of previously published research.

## Author Contributions

**Conceptualisation:** Pitak Benjarattanaporn, Peter Dodd, Debebe Adewo, Andrew Lee

**Data curation:** Pitak Benjarattanaporn

**Formal analysis:** Pitak Benjarattanaporn

**Methodology:** Pitak Benjarattanaporn, Peter Dodd, Debebe Adewo, Andrew Lee, Anthea Sutton

**Project administration:** Pitak Benjarattanaporn

**Resources:** Anthea Sutton

**Supervision:** Peter Dodd, Debebe Adewo, Andrew Lee

**Visualisation:** Pitak Benjarattanaporn

**Writing – original draft:** Pitak Benjarattanaporn

**Writing – review & editing:** Peter Dodd, Debebe Adewo, Andrew Lee, Anthea Sutton

## Acknowledgements

Pitak Benjarattanaporn, Peter Dodd, Debebe Adewo, Andrew Lee, and Anthea Sutton contributed to this study. The authors would like to thank Andrew Booth for his contribution to the framework selection.

TSW and KLT acknowledge funding provided through the UK–Southeast Asia Vaccine Manufacturing Research Hub. The Hub is funded by the Department of Health and Social Care using UK International Development funding and is managed by EPSRC. The views expressed in this publication are those of the author(s) and not necessarily those of the Department of Health and Social Care.

---

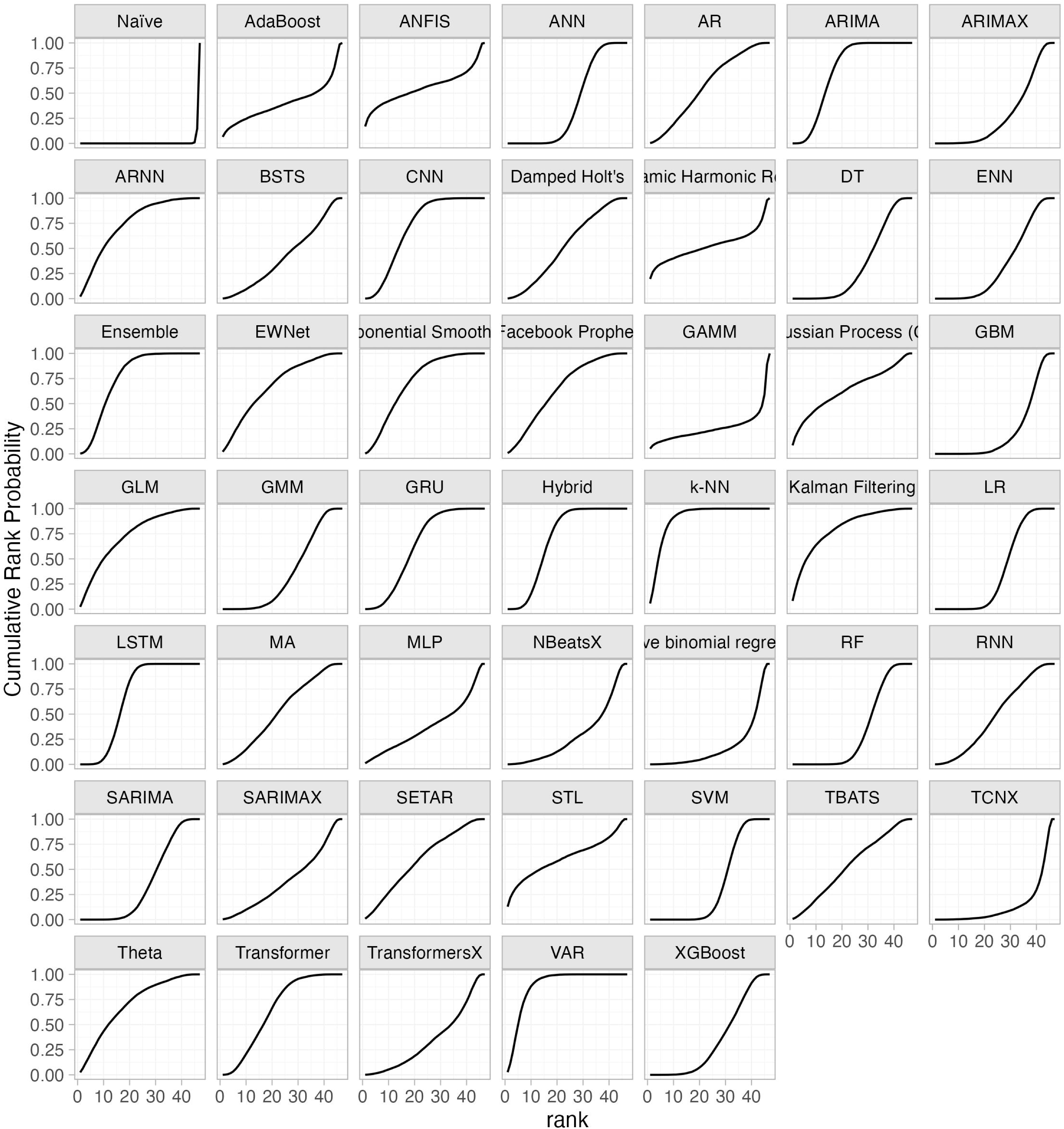

