## Supplementary Material 1: Appendices for "Dengue Forecasting Models: A Systematic Review and Network Meta-Analysis of Relative Performance"

### Appendix A: Search Strategies Methodology

#### Appendix Table A1: PICOS Framework

| | Included | Excluded |
|---|---|---|
| **Population (P)** | Populations/Settings that are susceptible to dengue outbreaks | Populations/Settings that are not susceptible to dengue outbreaks |
| **Intervention (I)** | Type(s) of forecasting methodology used to forecast dengue incidence.<br><br>This includes, but is not limited to, Statistical models, Machine learning models, Deep learning models, Hybrid models, and Combinations of multiple methods. | Compartmental models, Agent-Based Models (ABMs), and other non-forecasting predictive or projection methods. |
| **Comparator (C)** | Other forecasting models as defined in Intervention. | Other forecasting models as defined in Intervention. |
| **Outcome (O)** | The predictive performance of the models, as measured by quantitative evaluation metrics. | Outcomes related to the biological or epidemiological aspects of dengue |
| **Study Type (S)** | Comparative studies of two or more forecasting methodologies. | Studies that only present the development or application of a single forecasting model. |

### Appendix B: Search Strategy and List of Included Studies

#### Appendix B1: Search Strategy

##### MEDLINE Search Strategies

Ovid MEDLINE(R) Epub Ahead of Print and In-Process, In-Data-Review & Other Non-Indexed Citations and Daily <March 07, 2025>

1 Dengue/ or Dengue Virus/ 20163
2 dengue.ti,ab. 30691
3 DENV.ti,ab. 6968
4 1 or 2 or 3 32017
5 Forecasting/ 93567
6 forecast*.ti,ab. 34739
7 predict*.ti,ab. 2311352
8 5 or 6 or 7 2398793
9 4 and 8 2909
10 Models, Statistical/ 102043
11 Linear Models/ 88714
12 model*.ti,ab. 4248966
13 10 or 11 or 12 4341549
14 9 and 13 1254
15 limit 14 to yr="2014 -2025" 1037

##### Scopus Search Strategies

(TITLE-ABS-KEY(dengue) OR TITLE-ABS-KEY("dengue virus") OR TITLE-ABS-KEY(DENV))
AND
(TITLE-ABS-KEY(forecasting) OR TITLE-ABS-KEY(forecast*) OR TITLE-ABS-KEY(predict*))
AND
(TITLE-ABS-KEY("models, statistical") OR TITLE-ABS-KEY("linear models") OR TITLE-ABS-KEY(model*))
AND
(PUBYEAR > 2013 AND PUBYEAR < 2026)

#### Web of Science Search Strategies

(TS=(dengue) OR TS=("dengue virus") OR TS=(DENV))
AND
(TS=(forecasting) OR TS=(forecast*) OR TS=(predict*))
AND
(TS=("models, statistical") OR TS=("linear models") OR TS=(model*))
AND
PY=(2014-2025)

### Appendix Table B1: List of Included Studies

| Citation | Item Type | 1st Author and year | COUNTRY | Covariates | Covariates category | Incidence data granularity | Validation | CATEGORY MODELS | Metrics | Time Horizon |
|---|---|---|---|---|---|---|---|---|---|---|
| [1] | Conference Paper | Addawe, 2023 | Philippines | Incidence cases, Humidity, Rainfall, Temperature, Wind Speed | Epidemiological, Meteorological | Weekly | Other | Machine Learning | MAE, R^2 | 1M |
| [2] | Conference Paper | Agarwala, 2024 | Bangladesh | Incidence cases | Epidemiological | Daily | Unclear | Machine Learning, Time Series | RMSE, MAE, MSE, R^2 | 24M |
| [3] | Journal Article | Anggraeni, 2021 | Indonesia | Humidity, Incidence cases, Internet search query, Social Media Data, Rainfall, Temperature, Wind Speed | Meteorological, Epidemiological, and Internet Data | Not Specified | Cross-Validation | Machine Learning, Time Series, Deep Learning | RMSE, MAE, SMAPE | Not clear |
| [4] | Journal Article | Anggraeni, 2024 | Indonesia | Humidity, Incidence cases, Population, Mosquito density, Rainfall, Temperature, Wind Speed | Meteorological, Epidemiological, Demographic, Entomological | Weekly | Cross-Validation | Hybrid, Time Series, Machine Learning | MSE, MAPE | 1M,3M |
| [5] | Journal Article | Appice, 2020 | Mexico | Incidence cases, Temperature | Epidemiological, Meteorological | Monthly | Data Splitting | Machine Learning, Time Series | RMSE | 12M |

| | | | | | | | | | | |
|---|---|---|---|---|---|---|---|---|---|---|
| [6] | Conference Paper | Baker, 2021 | Peru, Puerto Rico | Humidity, Vegetation Index, Incidence cases, Rainfall, Temperature, Wind Speed | Meteorological, Geospatial data, Epidemiological | Weekly | Data Splitting | Machine Learning, Time Series, Ensemble | MAE | Not clear |
| [7] | Journal Article | Baquero, 2018 | Brazil | Humidity, Incidence cases, Rainfall, Temperature | Meteorological, Epidemiological | Daily | Cross-Validation | Hybrid, Machine Learning, Deep Learning, Time Series | RMSE | 16M |
| [8] | Journal Article | Benedum, 2020 | Peru, Puerto Rico, Singapore | Humidity, Incidence cases, Population, Rainfall, Temperature | Meteorological, Epidemiological, Demographic | Weekly | Data Splitting | Time Series, Machine Learning | MAE | 4W,12W |
| [9] | Conference Paper | Bogado, 2021 | Paraguay | Humidity, Incidence cases, Rainfall, Temperature, Wind Speed | Meteorological, Epidemiological | Weekly | Data Splitting | Machine Learning, Deep Learning | RMSE | Weekly |
| [10] | Journal Article | Carvajal, 2018 | Philippines | Flood level, Incidence per n population, El Niño-Southern Oscillation (ENSO), Temperature, Wind Speed | Geospatial data, Epidemiological, Meteorological | Yearly | Data Splitting | Machine Learning, Time Series | RMSE, MAE | 12M |
| [11] | Journal Article | Chakraborty, 2019 | Puerto Rico, Peru, Philippines | Incidence cases, Incidence per n population | Epidemiological | Weekly, Monthly | Unclear | Machine Learning, Time Series ,Hybrid | RMSE, MAE, SMAPE | 6M,1Y |
| [12] | Journal Article | Chakraborty, 2020 | Singapore | Humidity, Incidence cases, Rainfall, Temperature | Meteorological, Epidemiological | Weekly | Cross-Validation | Time Series, Machine Learning | RMSE, MSE | Not clear |

| | | | | | | | | | | |
|---|---|---|---|---|---|---|---|---|---|---|
| [13] | Journal Article | Chumpu, 2019 | Thailand | Humidity, Incidence cases, Rainfall, Temperature, Wind Speed | Meteorological, Epidemiological | Weekly | Data Splitting | Time Series | RMSE, R^2 | 1Y |
| [14] | Journal Article | Dhaked, 2025 | India | Humidity, Incidence cases, Rainfall, Temperature, Wind Speed | Meteorological, Epidemiological | Monthly | Data Splitting | Machine Learning, Deep Learning | RMSE, MAE, MSE | Monthly |
| [15] | Journal Article | Doni, 2020 | India | Humidity, Death cases, Incidence cases, Population, Rainfall, Temperature | Meteorological, Epidemiological, Demographic | Not Specified | Data Splitting | Machine Learning ,Deep Learning | RMSE | 1M |
| [16] | Journal Article | Guo, 2017 | China | Humidity, Internet search query, Incidence cases, Temperature | Meteorological, Internet Data, Epidemiological | Weekly | Cross-Validation | Machine Learning | RMSE, R^2 | 12W |
| [17] | Journal Article | Guo, 2018 | China | Humidity, Age, Date of illness onset, Death cases, Gender, Internet search query, Social Media Data, Incidence cases, Rainfall, Temperature | Meteorological, Demographic, Epidemiological, Internet Data | Weekly | Future-Oriented | Machine Learning | RMSE, MAE, Pearson's Correlation | 1W,2W,Leave-One-Out Cross-Validation |
| [18] | Journal Article | Handari, 2021 | Indonesia | Humidity, Incidence cases, Rainfall, Temperature | Meteorological, Epidemiological | Weekly | Data Splitting | Deep Learning | RMSE | Weekly |
| [19] | Journal Article | Hasan, 2024 | Bangladesh | Incidence cases | Epidemiological | Monthly | Future-Oriented | Time Series | RMSE, MAE, MSE | 12m |

| | | | | | | | | | | |
|---|---|---|---|---|---|---|---|---|---|---|
| [20] | Conference Paper | Ho, 2015 | Malaysia | Incidence cases | Epidemiological | Weekly | Data Splitting | Time Series,Machine Learning | RMSE, MAE, MAPE, MASE | 1Y 2M |
| [21] | Journal Article | Jayashree, 2015 | India | Humidity, Rainfall, Incidence cases, Temperature | Meteorological, Epidemiological | Not Specified | Unclear | Time Series | RMSE, MAE, ME | 4Y |
| [22] | Journal Article | Juraphanthong, 2021 | Thailand | Humidity, Rainfall, Incidence cases, Temperature | Meteorological, Epidemiological | Monthly | Unclear | Time Series,Hybrid | RMSE, MAPE | Monthly |
| [23] | Journal Article | Kakarla, 2023 | India | Humidity, El Niño-Southern Oscillation (ENSO), Rainfall, Incidence cases, Soil moisture, Temperature, Wind Speed | Meteorological, Epidemiological | Monthly | Cross-Validation | Machine Learning,Deep Learning,Time Series | RMSE, R^2, R | 1M |
| [24] | Journal Article | Kerdprasop, 2020 | Thailand | Humidity, El Niño-Southern Oscillation (ENSO), Rainfall, Vegetation Index, Incidence cases, Temperature, Wind Speed | Meteorological, Geospatial data, Epidemiological | Monthly | Data Splitting | Deep Learning, Machine Learning | RMSE, MAE, CORR | 1Y |
| [25] | Conference Paper | Khaira, 2020 | Indonesia | Incidence cases | Epidemiological | Not Specified | Data Splitting | Deep Learning,Time Series | RMSE, MAE | 6M |
| [26] | Journal Article | Koplewitz, 2022 | Brazil | Humidity, Rainfall, Incidence cases, Internet search | Meteorological, Epidemiological, | Weekly | Future-Oriented | Machine Learning | RMSE, R^2, Pearson's Correlation | Nowcast |

| | | | | | | | | | | |
|---|---|---|---|---|---|---|---|---|---|---|
| | | | | query, Wind Speed | Internet Data | | | | | |
| [27] | Journal Article | Li, 2022 | Brazil | Humidity, Rainfall, Vegetation Index, Incidence cases, Global Artificial Impervious Area (GAIA), Temperature | Meteorological, Geospatial data, Epidemiological | Weekly | Data Splitting | Deep Learning, Machine Learning | RMSE, MAE | 1W,2W,3W,4W |
| [28] | Journal Article | Ligue, 2022 | Philippines | Humidity, Rainfall, Incidence cases, Temperature, Wind Speed | Meteorological, Epidemiological | Monthly | Data Splitting | Time Series, Deep Learning | RMSE | Monthly |
| [29] | Journal Article | Lima, 2020 | Brazil | Incidence cases | Epidemiological | Monthly | Data Splitting | Time Series,Machine Learning,Deep Learning, Hybrid | MAPE | 3M,6M |
| [30] | Journal Article | Liu, 2019 | China | Humidity, Rainfall, Age, Gender, Death cases, Internet search query, Incidence cases, Occupation, Address, Temperature | Meteorological, Demographic, Epidemiological, Internet Data, Geospatial data | Monthly | Data Splitting | Machine Learning | RMSE, R^2 | 7M |
| [31] | Journal Article | Mahdiana, 2017 | Indonesia | Humidity, Rainfall, Population, Incidence cases, Temperature | Meteorological, Demographic, Epidemiological | Monthly | Unclear | Time Series,Machine Learning | RMSE, MAE | 12M |

| | | | | | | | | | | |
|---|---|---|---|---|---|---|---|---|---|---|
| [32] | Journal Article | Majeed, 2023 | Malaysia | Humidity, Rainfall, Water bodies, Vegetation Index, Road networks, Incidence cases, Population, Land cover, Elevation, Temperature, Wind Speed | Meteorological, Geospatial data, Epidemiological, Demographic | Weekly | Data Splitting | Machine Learning | RMSE | 3M |
| [33] | Conference Paper | Munarsih, 2020 | Indonesia | Incidence cases | Epidemiological | Not Specified | Unclear | Time Series,Machine Learning | MAE, MSE | Not clear |
| [34] | Journal Article | Mustaffa, 2024 | Malaysia | Incidence cases | Epidemiological | Weekly | Data Splitting | Time Series,Hybrid | RMSE, MAPE | Weekly |
| [35] | Journal Article | Nabilah, 2023 | Malaysia | Humidity, Rainfall, Incidence cases, Temperature, Wind Speed | Meteorological, Epidemiological | Monthly | Data Splitting | Machine Learning, Ensemble | RMSE, MSE, SMAPE | Weekly |
| [36] | Journal Article | Navarro Valencia, 2021 | Panama | Humidity, Rainfall, Incidence cases, Temperature | Meteorological, Epidemiological | Weekly | Data Splitting | Deep Learning,Time Series | RMSE, MAPE | 1Y |
| [37] | Journal Article | Nguyen, 2022 | Vietnam | Humidity, Rainfall, Incidence cases, Temperature | Meteorological, Epidemiological | Monthly | Data Splitting | Machine Learning,Deep Learning,Time Series | RMSE, MAE | Monthly |
| [38] | Journal Article | Othman, 2022 | Indonesia | Incidence cases | Epidemiological | Monthly | Data Splitting | Time Series,Deep Learning | RMSE, MAE, MAPE | 1M |

| | | | | | | | | | | |
|---|---|---|---|---|---|---|---|---|---|---|
| [39] | Journal Article | Panja, 2023 | India, Puerto Rico, Peru | Rainfall, Incidence cases | Meteorological, Epidemiological | Weekly | Data Splitting | Machine Learning, Time Series, Hybrid, Ensemble, Deep Learning | RMSE, MAE, MASE, SMAPE | 26W,52W |
| [40] | Journal Article | Patil, 2021 | India | Humidity, Rainfall, Incidence cases, Temperature | Meteorological, Epidemiological | Not Specified | Future-Oriented | Time Series, Machine Learning | RMSE, MAE, R^2 | 3Y |
| [41] | Journal Article | Patra, 2024 | Laos | Incidence cases | Epidemiological | Weekly | Data Splitting | Machine Learning, Deep Learning | RMSE, MAE, R | 1W |
| [42] | Conference Paper | Pham, 2018 | Malaysia | Humidity, Rainfall, Vegetation Index, Incidence cases, Temperature, Wind Speed | Meteorological, Geospatial data, Epidemiological | Weekly | Data Splitting | Hybrid, Machine Learning | RMSE, MAE | 2W |
| [43] | Journal Article | Phung, 2014 | Vietnam | Rainfall, Incidence cases, Temperature | Meteorological, Epidemiological | Monthly | Data Splitting | Machine Learning, Time Series | MAPE | 3M,6M,9M,12M |
| [44] | Journal Article | Polwiang, 2020 | Thailand | Humidity, Rainfall, Incidence cases, Temperature | Meteorological, Epidemiological | Monthly | Data Splitting | Machine Learning, Time Series | RMSE, MAE, MAPE, CORR | 1Y |
| [45] | Conference Paper | Prome, 2024 | Bangladesh | Humidity, Rainfall, Incidence cases, Vegetation Index, Temperature, Wind Speed | Meteorological, Epidemiological, Geospatial data | Monthly | Data Splitting | Machine Learning | RMSE, MAE, MSE, R^2 | Not clear |

| | | | | | | | | | | |
|---|---|---|---|---|---|---|---|---|---|---|
| [46] | Journal Article | Puengpreeda, 2020 | Thailand | Rainfall, Incidence cases, Internet search query, Temperature | Meteorological, Epidemiological, Internet Data | Monthly | Data Splitting | Machine Learning, Time Series | MAE, MSE, R^2 | 1W,2W,3W,4W |
| [47] | Conference Paper | Rajendran, 2023 | India | Humidity, Rainfall, Incidence cases, Temperature | Meteorological, Epidemiological | Monthly | Data Splitting | Deep Learning, Machine Learning | MAE | 1M |
| [48] | Journal Article | Rangarajan, 2019 | Brazil, Mexico, Singapore, Taiwan, Thailand | Incidence cases, Internet search query | Epidemiological, Internet Data | Monthly, Weekly | Cross-Validation | Machine Learning, Ensemble, Time Series, Hybrid | RMSE, MAE, MAPE | Real-time |
| [49] | Journal Article | Sebastianelli, 2024 | Brazil | Humidity, Rainfall, Vegetation Index, Age, Population, Temperature, Socio-economic data, Elevation, Incidence per n population, Wind Speed | Meteorological, Geospatial data, Demographic, Socio-economic data, Epidemiological | Monthly | Data Splitting | Machine Learning, Ensemble, Deep Learning | nRMSE | Monthly |
| [50] | Journal Article | Sharma, 2021 | Hongkok | Incidence cases | Epidemiological | Monthly | Future-Oriented | Time Series, Machine Learning, Hybrid | MAE | Not clear |
| [51] | Journal Article | Shashvat, 2019 | India | Humidity, Rainfall, Incidence cases | Meteorological, Epidemiological | Monthly | Unclear | Machine Learning, Ensemble, Time Series | RMSE, MAE, MSE | 12M |
| [52] | Conference Paper | Shashvat, 2023 | India | Incidence cases | Epidemiological | Monthly | Data Splitting | Machine Learning, Ensemble | RMSE, MAE, MSE | 12M |

| | | | | | | | | | | |
|---|---|---|---|---|---|---|---|---|---|---|
| [53] | Journal Article | Shi, 2015 | Singapore | Humidity, Temperature, Mosquito density, Population, Incidence cases | Meteorological, Entomological, Demographic, Epidemiological | Weekly | Cross-Validation | Machine Learning, Time Series | MAPE | 3M |
| [54] | Journal Article | Tian, 2024 | Singapore | Humidity, Rainfall, Temperature, Incidence cases | Meteorological, Epidemiological | Weekly | Cross-Validation | Machine Learning | RMSE, MAE, R | Weekly |
| [55] | Journal Article | Tuan, 2024 | Vietnam | Humidity, Rainfall, Temperature, Incidence cases, Wind Speed | Meteorological, Epidemiological | Weekly | Cross-Validation | Machine Learning, Deep Learning, Time Series | RMSE, MAE | Monthly |
| [56] | Conference Paper | Weng, 2024 | Sri Lanka | Humidity, Rainfall, Temperature, Land cover, Incidence cases, Wind Speed | Meteorological, Geospatial data, Epidemiological | Weekly | Cross-Validation | Time Series, Deep Learning, Machine Learning | RMSE, MAE | 3W |
| [57] | Journal Article | Xu, 2020 | China | Humidity, Rainfall, Temperature, Incidence cases, Wind Speed | Meteorological, Epidemiological | Monthly | Data Splitting | Machine Learning, Deep Learning | RMSE, MAE | 24M |
| [58] | Journal Article | Zhao, 2020 | Colombia | Rainfall, Temperature, Vegetation Index, Incidence cases, Population, Gini index, Education Coverage | Meteorological, Geospatial data, Epidemiological, Demographic, Socio-economic data | Weekly | Cross-Validation | Machine Learning | RMSE, MAE | 1W,2W,3W,4W,5W,6W,7W,8W,9W,10W,11W,12W |

| | | | | | | | | | | |
|---|---|---|---|---|---|---|---|---|---|---|
| [59] | Journal Article | Zhao, 2023 | Singapore | Humidity, Rainfall, Temperature, Incidence cases | Meteorological, Epidemiological | Weekly | Data Splitting | Deep Learning, Ensemble, Hybrid | RMSE, MAE, MAPE | 1W,2W,3W,4W |

#### Appendix Table B2: Distribution of Forecasting Categories and Specific Model Implementations across Included Studies

| Methodology Category | Specific Model | Study Count (n =59) | Total Usage Count | References |
|---|---|---|---|---|
| **Deep Learning** | ANFIS | 1 | 1 | [24] |
| | ARNN | 2 | 8 | [39,56] |
| | CNN | 4 | 30 | [14,37,41,59] |
| | ENN | 1 | 30 | [18] |
| | EWNet | 1 | 6 | [39] |
| | GRU | 2 | 38 | [18,59] |
| | LSTM | 19 | 233 | [3,9,14,15,18,23,25,27,28,36–38,41,47,49,55–57,59] |
| | MLP | 4 | 18 | [7,24,28,29] |
| | NBeatsX | 1 | 6 | [39] |
| | RNN | 2 | 14 | [39,59] |
| | Transformer | 1 | 20 | [37] |
| **Ensemble** | ANN + LR + SVm | 1 | 1 | [51] |
| | ARIMA-WARIMA | 1 | 6 | [39] |
| | CatBoost+SVM+LSTM | 1 | 27 | [49] |
| | CNN + LSTM | 1 | 8 | [59] |
| | Holt-Winters Based | 1 | 10 | [48] |
| | RF + DT | 1 | 1 | [6] |
| | RF + NB + DT + SVM | 1 | 1 | [6] |
| | SCAD + Elastic Net[35] | 1 | 1 | [35] |
| | SCAD + Elastic Net + Ridge | 1 | 1 | [35] |
| | SCAD + Elastic Net + Ridge + MCP | 1 | 1 | [35] |
| | SCAD + Elastic Net + Ridge + MCP + LASSO | 1 | 1 | [35] |
| | SVM + LR + ANN | 1 | 1 | [52] |

| | | | | |
|---|---|---|---|---|
| | XEWNet | 1 | 6 | [39] |
| **Hybrid** | ARIMA - ANN | 1 | 6 | [4] |
| | ARIMA - ARNN | 1 | 6 | [39] |
| | ARIMA - SVM | 1 | 6 | [4] |
| | ARIMA - ANN | 1 | 3 | [11] |
| | ARIMA - ETS - Theta | 1 | 6 | [39] |
| | ARIMA - NNAR | 1 | 3 | [11] |
| | ARIMAS | 1 | 1 | [22] |
| | ARNN - Theta - ETS | 1 | 6 | [39] |
| | ARNNX | 1 | 6 | [39] |
| | CNN - BiLSTM | 1 | 24 | [59] |
| | CNN - GRU | 1 | 24 | [59] |
| | CNN - LSTM | 1 | 24 | [59] |
| | CNN - RNN | 1 | 24 | [59] |
| | EMD - GRNN - PSO | 1 | 6 | [4] |
| | GA - RNN | 1 | 1 | [42] |
| | GAM - ANN - SARIMA. | 1 | 1 | [7] |
| | NNAR | 3 | 5 | [11,34,50] |
| | NNETAR | 1 | 9 | [29] |
| **Machine Learning** | AdaBoost | 3 | 6 | [6,45,46] |
| | ANN | 14 | 144 | [3,5,6,11,14,15,24,32,39,44,51,52,57,58] |
| | CatBoost | 1 | 27 | [49] |
| | DT | 6 | 39 | [6,24,32,40,45,54] |
| | ELM | 1 | 27 | [29] |
| | Exponential Smoothing | 5 | 30 | [20,29,33,39,50] |
| | GAMM | 1 | 1 | [30] |
| | Gaussian Process (GP) | 2 | 2 | [12,45] |
| | GBM | 6 | 33 | [1,10,16,23,54,57] |

| | | | | |
|---|---|---|---|---|
| | GLM | 4 | 11 | [24,43,48,54] |
| | GMM | 8 | 39 | [4,7,10,12,15,16,30,57] |
| | GRNN | 1 | 6 | [4] |
| | k-NN | 2 | 33 | [5,6] |
| | LR | 21 | 149 | [1,3,4,6,8,9,16,17,26,31,35,37,40,42–45,51–53,55] |
| | NBM (negative binomial regression model) | 1 | 6 | [16] |
| | RF | 17 | 112 | [1,2,5,6,8–10,12,26,27,40,45,46,50,54,56,58] |
| | SVM | 17 | 224 | [3,5,6,9,15,16,23,24,32,37,40,45,47,49,51,52,54,57] |
| | XGBoost | 7 | 28 | [2,15,37,45,54–56] |
| **Time Series** | AR | 1 | 9 | [40] |
| | ARIMA | 20 | 124 | [5,8,11–13,19–22,28,29,31,38–40,40,44,50,51,56] |
| | ARIMAX | 4 | 29 | [3,4,22,39] |
| | BATS | 1 | 12 | [29] |
| | BSTS | 1 | 6 | [39] |
| | Damped Holt's | 3 | 13 | [20,40,50] |
| | DHR | 1 | 1 | [34] |
| | Facebook Prophet | 1 | 9 | [40] |
| | Kalman Filtering | 2 | 11 | [4,48] |
| | MA | 3 | 26 | [40,46,50] |
| | Naïve | 6 | 48 | [6,7,29,46,48,50] |
| | SARIMA | 15 | 56 | [2,7,13,19–21,25,28,34,36–38,40,43,53] |
| | SARIMAX | 4 | 11 | [4,10,36,55] |
| | SETAR | 1 | 6 | [39] |
| | STL | 1 | 2 | [20] |

| | | | |
|---|---|---|---|
| StructTS | 1 | 14 | [29] |
| TBATS | 2 | 18 | [29,39] |
| TCNX | 1 | 6 | [39] |
| Theta | 2 | 7 | [39,50] |
| TransformersX | 1 | 6 | [39] |
| VAR | 3 | 34 | [5,23,31] |
| | **Total** | **1995** | |

### Appendix C: Quantitative Synthesis Supplementary Data

#### Appendix Table C1: NMA Summary Ranking

| Model* | SCURA Value | Mean Rank | SD | 95% CrI (Lower, Upper) | Bulk_ESS | Tail_ESS | Rhat |
|---|---|---|---|---|---|---|---|
| k-NN | 90.26% | -2.32 | 0.40 | (-3.08, -1.52) | 250.20 | 506.51 | 1.012 |
| VAR | 88.44% | -2.29 | 0.39 | (-3.04, -1.52) | 250.55 | 575.38 | 1.012 |
| Kalman Filtering | 79.08% | -2.23 | 0.44 | (-3.10, -1.36) | 382.84 | 1,027.71 | 1.009 |
| GLM | 72.94% | -2.14 | 0.42 | (-2.96, -1.29) | 348.91 | 815.72 | 1.008 |
| ARNN | 74.13% | -2.14 | 0.43 | (-2.97, -1.28) | 300.86 | 714.94 | 1.010 |
| Ensemble | 75.88% | -2.12 | 0.38 | (-2.86, -1.35) | 243.18 | 553.96 | 1.012 |
| Theta | 69.83% | -2.10 | 0.44 | (-2.95, -1.22) | 317.87 | 849.44 | 1.009 |
| Exponential Smoothing | 70.93% | -2.09 | 0.41 | (-2.87, -1.28) | 273.71 | 710.25 | 1.011 |
| EWNet | 67.99% | -2.08 | 0.45 | (-2.93, -1.19) | 318.70 | 780.26 | 1.010 |
| Gaussian Process | 63.29% | -2.08 | 0.56 | (-3.17, -0.98) | 498.04 | 1,409.81 | 1.006 |
| STL | 61.57% | -2.08 | 0.65 | (-3.34, -0.79) | 710.59 | 2,015.10 | 1.005 |
| ARIMA | 70.90% | -2.07 | 0.39 | (-2.79, -1.31) | 234.83 | 525.21 | 1.013 |
| CNN | 69.94% | -2.07 | 0.39 | (-2.82, -1.28) | 248.08 | 545.41 | 1.013 |
| Hybrid | 68.62% | -2.05 | 0.38 | (-2.78, -1.31) | 235.27 | 525.44 | 1.013 |
| Facebook Prophet | 64.48% | -2.04 | 0.43 | (-2.87, -1.18) | 284.12 | 749.90 | 1.011 |
| LSTM | 65.61% | -2.02 | 0.38 | (-2.75, -1.28) | 232.22 | 469.07 | 1.013 |
| Transformer | 64.41% | -2.02 | 0.40 | (-2.81, -1.25) | 250.01 | 574.33 | 1.013 |
| GRU | 62.34% | -2.00 | 0.39 | (-2.77, -1.22) | 243.48 | 553.24 | 1.012 |
| SETAR | 59.43% | -2.00 | 0.45 | (-2.87, -1.12) | 306.40 | 756.69 | 1.010 |
| ANFIS | 54.93% | -1.99 | 0.82 | (-3.60, -0.42) | 1,139.80 | 3,371.28 | 1.002 |
| DHR | 53.06% | -1.96 | 0.95 | (-3.81, -0.11) | 1,346.75 | 3,915.19 | 1.002 |
| AR | 56.56% | -1.96 | 0.43 | (-2.79, -1.10) | 278.88 | 668.32 | 1.011 |
| TBATS | 54.04% | -1.94 | 0.45 | (-2.81, -1.07) | 324.78 | 698.72 | 1.009 |
| Damped Holt's | 52.55% | -1.93 | 0.42 | (-2.73, -1.09) | 281.95 | 680.31 | 1.012 |
| MA | 52.47% | -1.93 | 0.43 | (-2.78, -1.08) | 292.21 | 735.76 | 1.010 |
| RNN | 46.89% | -1.88 | 0.41 | (-2.67, -1.07) | 271.92 | 704.47 | 1.011 |
| BSTS | 41.65% | -1.83 | 0.45 | (-2.68, -0.93) | 327.45 | 922.18 | 1.009 |
| LR | 37.87% | -1.81 | 0.38 | (-2.54, -1.06) | 232.88 | 520.19 | 1.014 |
| ANN | 37.93% | -1.81 | 0.38 | (-2.55, -1.06) | 235.25 | 479.36 | 1.013 |
| SARIMA | 35.54% | -1.79 | 0.39 | (-2.54, -1.03) | 240.70 | 572.51 | 1.013 |
| SVM | 34.03% | -1.79 | 0.38 | (-2.52, -1.04) | 231.55 | 501.86 | 1.013 |
| GMM | 34.76% | -1.78 | 0.39 | (-2.54, -1.02) | 246.32 | 544.96 | 1.013 |
| RF | 32.98% | -1.78 | 0.39 | (-2.51, -1.03) | 235.30 | 547.22 | 1.013 |

| | | | | | | | |
|---|---|---|---|---|---|---|---|
| ENN | 33.28% | -1.77 | 0.40 | (-2.55, -0.97) | 245.03 | 549.75 | 1.012 |
| XGBoost | 33.24% | -1.77 | 0.39 | (-2.53, -0.99) | 245.59 | 490.55 | 1.012 |
| SARIMAX | 37.35% | -1.77 | 0.47 | (-2.67, -0.84) | 346.40 | 924.65 | 1.008 |
| DT | 32.72% | -1.77 | 0.39 | (-2.52, -1.00) | 244.58 | 563.04 | 1.012 |
| TransformersX | 34.09% | -1.75 | 0.45 | (-2.60, -0.87) | 317.76 | 910.86 | 1.010 |
| MLP | 36.84% | -1.74 | 0.50 | (-2.70, -0.73) | 503.34 | 1,453.59 | 1.005 |
| ARIMAX | 28.28% | -1.72 | 0.40 | (-2.50, -0.93) | 261.51 | 568.18 | 1.011 |
| AdaBoost | 40.15% | -1.71 | 0.76 | (-3.22, -0.22) | 890.17 | 3,026.64 | 1.003 |
| NBeatsX | 28.52% | -1.68 | 0.45 | (-2.56, -0.80) | 314.78 | 733.44 | 1.009 |
| GBM | 22.43% | -1.67 | 0.40 | (-2.44, -0.90) | 245.17 | 545.83 | 1.012 |
| NBM | 17.74% | -1.53 | 0.46 | (-2.41, -0.63) | 318.64 | 762.34 | 1.010 |
| TCNX | 14.76% | -1.47 | 0.45 | (-2.36, -0.57) | 326.66 | 859.92 | 1.009 |
| GAMM | 24.95% | -1.23 | 0.96 | (-3.12, 0.66) | 1,387.74 | 3,670.73 | 1.001 |

*** Reference Model: Naïve**

### Appendix Figure C1: Cumulative Rank Probability Curves

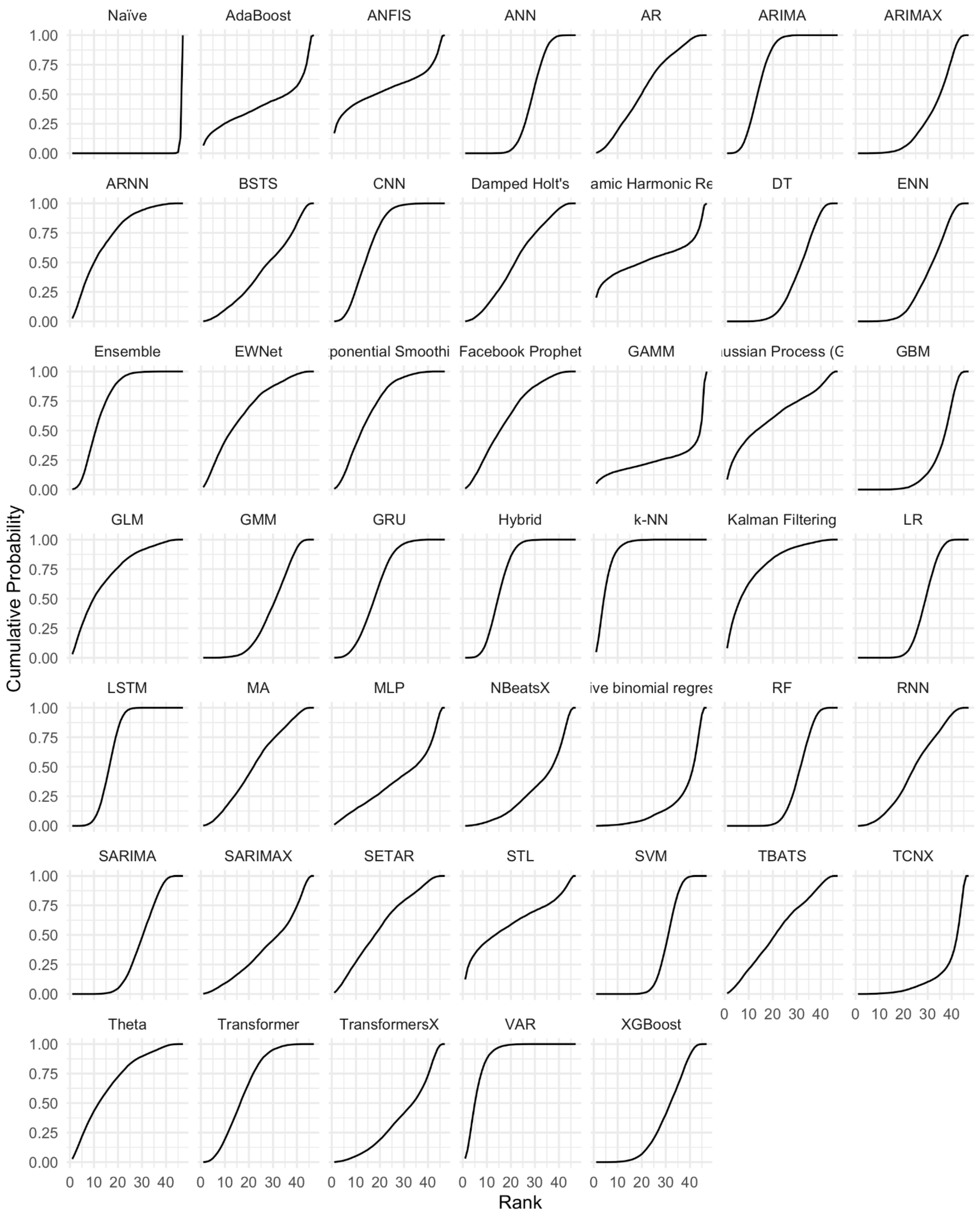

### Appendix Table C2: Evaluation Metrics

| # | *First author, year* | RMSE | MAE | ME | MSE | nRMSE | MAPE | MASE | SMAPE | R-Squa | R | CORR | ρ |
|---|---|---|---|---|---|---|---|---|---|---|---|---|---|
| 1 | Addawe, 2023 | 0 | 1 | 0 | 0 | 0 | 0 | 0 | 0 | 1 | 0 | 0 | 0 |
| 2 | Agarwala, 2024 | 1 | 1 | 0 | 1 | 0 | 0 | 0 | 0 | 1 | 0 | 0 | 0 |
| 3 | Anggraeni, 2021 | 1 | 1 | 0 | 0 | 0 | 0 | 0 | 1 | 0 | 0 | 0 | 0 |
| 4 | Anggraeni, 2024 | 0 | 0 | 0 | 1 | 0 | 1 | 0 | 0 | 0 | 0 | 0 | 0 |
| 5 | Appice, 2020 | 1 | 0 | 0 | 0 | 0 | 0 | 0 | 0 | 0 | 0 | 0 | 0 |
| 6 | Baker, 2021 | 0 | 1 | 0 | 0 | 0 | 0 | 0 | 0 | 0 | 0 | 0 | 0 |
| 7 | Baquero, 2018 | 1 | 0 | 0 | 0 | 0 | 0 | 0 | 0 | 0 | 0 | 0 | 0 |
| 8 | Benedum, 2020 | 0 | 1 | 0 | 0 | 0 | 0 | 0 | 0 | 0 | 0 | 0 | 0 |
| 9 | Bogado, 2021 | 1 | 0 | 0 | 0 | 0 | 0 | 0 | 0 | 0 | 0 | 0 | 0 |
| 10 | Carvajal, 2018 | 1 | 1 | 0 | 0 | 0 | 0 | 0 | 0 | 0 | 0 | 0 | 0 |
| 11 | Chakraborty, 2019 | 1 | 1 | 0 | 0 | 0 | 0 | 0 | 1 | 0 | 0 | 0 | 0 |
| 12 | Chakraborty, 2020 | 1 | 0 | 0 | 1 | 0 | 0 | 0 | 0 | 0 | 0 | 0 | 0 |
| 13 | Chumpu, 2019 | 1 | 0 | 0 | 0 | 0 | 0 | 0 | 0 | 1 | 0 | 0 | 0 |
| 14 | Dhaked, 2025 | 1 | 1 | 0 | 1 | 0 | 0 | 0 | 0 | 0 | 0 | 0 | 0 |
| 15 | Doni, 2020 | 1 | 0 | 0 | 0 | 0 | 0 | 0 | 0 | 0 | 0 | 0 | 0 |
| 16 | Guo, 2017 | 1 | 0 | 0 | 0 | 0 | 0 | 0 | 0 | 1 | 0 | 0 | 0 |
| 17 | Guo, 2018 | 1 | 1 | 0 | 0 | 0 | 0 | 0 | 0 | 0 | 0 | 0 | 1 |
| 18 | Handari, 2021 | 1 | 0 | 0 | 0 | 0 | 0 | 0 | 0 | 0 | 0 | 0 | 0 |
| 19 | Hasan, 2024 | 1 | 1 | 0 | 1 | 0 | 0 | 0 | 0 | 0 | 0 | 0 | 0 |
| 20 | Ho, 2015 | 1 | 1 | 0 | 0 | 0 | 1 | 1 | 0 | 0 | 0 | 0 | 0 |
| 21 | Jayashree, 2015 | 1 | 1 | 1 | 0 | 0 | 0 | 0 | 0 | 0 | 0 | 0 | 0 |
| 22 | Juraphanthong, 2021 | 1 | 0 | 0 | 0 | 0 | 1 | 0 | 0 | 0 | 0 | 0 | 0 |
| 23 | Kakarla, 2023 | 1 | 0 | 0 | 0 | 0 | 0 | 0 | 0 | 1 | 1 | 0 | 0 |
| 24 | Kerdprasop, 2020 | 1 | 1 | 0 | 0 | 0 | 0 | 0 | 0 | 0 | 0 | 1 | 0 |
| 25 | Khaira, 2020 | 1 | 1 | 0 | 0 | 0 | 0 | 0 | 0 | 0 | 0 | 0 | 0 |
| 26 | Koplewitz, 2022 | 1 | 0 | 0 | 0 | 0 | 0 | 0 | 0 | 1 | 0 | 0 | 1 |
| 27 | Li, 2022 | 1 | 1 | 0 | 0 | 0 | 0 | 0 | 0 | 0 | 0 | 0 | 0 |
| 28 | Ligue, 2022 | 1 | 0 | 0 | 0 | 0 | 0 | 0 | 0 | 0 | 0 | 0 | 0 |

(continued from previous page)

| # | *First author, year* | RMSE | MAE | ME | MSE | nRMSE | MAPE | MASE | SMAPE | R-Squa | R | CORR | ρ |
|---|---|---|---|---|---|---|---|---|---|---|---|---|---|
| 29 | Lima, 2020 | 0 | 0 | 0 | 0 | 0 | 1 | 0 | 0 | 0 | 0 | 0 | 0 |
| 30 | Liu, 2019 | 1 | 0 | 0 | 0 | 0 | 0 | 0 | 0 | 1 | 0 | 0 | 0 |
| 31 | Mahdiana, 2017 | 1 | 1 | 0 | 0 | 0 | 0 | 0 | 0 | 0 | 0 | 0 | 0 |
| 32 | Majeed, 2023 | 1 | 0 | 0 | 0 | 0 | 0 | 0 | 0 | 0 | 0 | 0 | 0 |
| 33 | Munarsih, 2020 | 0 | 1 | 0 | 1 | 0 | 0 | 0 | 0 | 0 | 0 | 0 | 0 |
| 34 | Mustaffa, 2024 | 1 | 0 | 0 | 0 | 0 | 1 | 0 | 0 | 0 | 0 | 0 | 0 |
| 35 | Nabilah, 2023 | 1 | 0 | 0 | 1 | 0 | 0 | 0 | 1 | 0 | 0 | 0 | 0 |
| 36 | Navarro Valencia, 2021 | 1 | 0 | 0 | 0 | 0 | 1 | 0 | 0 | 0 | 0 | 0 | 0 |
| 37 | Nguyen, 2022 | 1 | 1 | 0 | 0 | 0 | 0 | 0 | 0 | 0 | 0 | 0 | 0 |
| 38 | Othman, 2022 | 1 | 1 | 0 | 0 | 0 | 1 | 0 | 0 | 0 | 0 | 0 | 0 |
| 39 | Panja, 2023 | 1 | 1 | 0 | 0 | 0 | 0 | 1 | 1 | 0 | 0 | 0 | 0 |
| 40 | Patil, 2021 | 1 | 1 | 0 | 0 | 0 | 0 | 0 | 0 | 1 | 0 | 0 | 0 |
| 41 | Patra, 2024 | 1 | 1 | 0 | 0 | 0 | 0 | 0 | 0 | 0 | 1 | 0 | 0 |
| 42 | Pham, 2018 | 1 | 1 | 0 | 0 | 0 | 0 | 0 | 0 | 0 | 0 | 0 | 0 |
| 43 | Phung, 2014 | 0 | 0 | 0 | 0 | 0 | 1 | 0 | 0 | 0 | 0 | 0 | 0 |
| 44 | Polwiang, 2020 | 1 | 1 | 0 | 0 | 0 | 1 | 0 | 0 | 0 | 0 | 1 | 0 |
| 45 | Prome, 2024 | 1 | 1 | 0 | 1 | 0 | 0 | 0 | 0 | 1 | 0 | 0 | 0 |
| 46 | Puengpreeda, 2020 | 0 | 1 | 0 | 1 | 0 | 0 | 0 | 0 | 1 | 0 | 0 | 0 |
| 47 | Rajendran, 2023 | 0 | 1 | 0 | 0 | 0 | 0 | 0 | 0 | 0 | 0 | 0 | 0 |
| 48 | Rangarajan, 2019 | 1 | 1 | 0 | 0 | 0 | 1 | 0 | 0 | 0 | 0 | 0 | 0 |
| 49 | Sebastianelli, 2024 | 0 | 0 | 0 | 0 | 1 | 0 | 0 | 0 | 0 | 0 | 0 | 0 |
| 50 | Sharma, 2021 | 0 | 1 | 0 | 0 | 0 | 0 | 0 | 0 | 0 | 0 | 0 | 0 |
| 51 | Shashvat, 2019 | 1 | 1 | 0 | 1 | 0 | 0 | 0 | 0 | 0 | 0 | 0 | 0 |
| 52 | Shashvat, 2023 | 1 | 1 | 0 | 1 | 0 | 0 | 0 | 0 | 0 | 0 | 0 | 0 |
| 53 | Shi, 2015 | 0 | 0 | 0 | 0 | 0 | 1 | 0 | 0 | 0 | 0 | 0 | 0 |
| 54 | Tian, 2024 | 1 | 1 | 0 | 0 | 0 | 0 | 0 | 0 | 0 | 1 | 0 | 0 |
| 55 | Tuan, 2024 | 1 | 1 | 0 | 0 | 0 | 0 | 0 | 0 | 0 | 0 | 0 | 0 |
| 56 | Weng, 2024 | 1 | 1 | 0 | 0 | 0 | 0 | 0 | 0 | 0 | 0 | 0 | 0 |
| 57 | Xu, 2020 | 1 | 1 | 0 | 0 | 0 | 0 | 0 | 0 | 0 | 0 | 0 | 0 |
| 58 | Zhao, 2020 | 1 | 1 | 0 | 0 | 0 | 0 | 0 | 0 | 0 | 0 | 0 | 0 |
| 59 | Zhao, 2023 | 1 | 1 | 0 | 0 | 0 | 1 | 0 | 0 | 0 | 0 | 0 | 0 |
| 60 | **Total** | **47** | **37** | **1** | **11** | **1** | **12** | **2** | **4** | **10** | **3** | **2** | **2** |

### Appendix D: Assessment Details

#### Appendix Table D1: Operational Definitions for Qualitative Scoring

| Dimension & Goal | Definition | Score | Operational Definition (Technical/Modeller Focus) |
|---|---|---|---|
| **Availability of open-source Python/R packages** | The degree to which a forecasting model's implementation is accessible and reproducible for public health analysts is typically measured by its availability in standardised, open-source programming packages. | **Very High** | Model is implemented and maintained in dedicated, language-agnostic open-source libraries (e.g., Python/R packages like EpiModel or forecast). |
| | | **High** | Core model logic is available in a widely used programming environment (e.g., R code snippet, Matlab function) but may not be fully packaged. |
| | | **Moderate** | The algorithm is well-described in the literature, but requires significant custom coding/reimplementation by the modeller. |
| | | **Low** | No code is provided; the algorithm is complex and requires full reproduction from the ground up, or is proprietary/commercial. |
| **Transparency/Interpretability** | The ability to clearly understand and communicate the relationship between the input covariates (e.g., climate data) and the resulting dengue prediction is critical for policy adoption and public health messaging. | **Very High** | Provides explicit, quantitative coefficients (e.g., regression weights) or directly derived parameters that explain the relationship between covariates and prediction. |
| | | **High** | Provides clear visualisation of feature importance (e.g., VAR/ARIMA coefficients for time lags) or uses model-agnostic techniques (e.g., SHAP, LIME) to explain local predictions. |
| | | **Moderate** | Interpretation relies on complex visualisation of decision boundaries or feature interactions; the core prediction mechanism remains mostly opaque. |
| | | **Low** | Prediction is derived from highly non-linear, deep mechanisms (e.g., deep neural networks) with no practical way to explain the underlying logic to a non-expert. |
| **Complexity of Parameter Tuning** | The technical effort and specialised knowledge required to calibrate a model's parameters to achieve optimal predictive performance for a specific dengue dataset. | **Very High** | Requires extensive hyperparameter optimisation (e.g., Deep Learning models); tuning is highly dependent on proprietary infrastructure. |
| | | **High** | Requires sophisticated methods (e.g., cross-validation grid search, nested-cross-validation) to set multiple non-intuitive parameters (e.g., hidden layers, regularisation). |
| | | **Moderate** | Parameters are few (2-4) and can be set based on common practice or simple fitting routines (e.g., AIC/BIC, lag selection). |
| | | **Low** | No explicit tuning is required; parameters are estimated directly from the data using standard, documented statistical procedures (e.g., Maximum Likelihood). |
| **Computational Requirements** | The necessary hardware (e.g., CPU, GPU, RAM) and time investment required to train the model and generate a prediction (inference), reflecting the cost and feasibility for routine public health deployment. | **Very High** | Training requires dedicated cloud computing resources (e.g., multiple GPUs) or HPC infrastructure; inference time (prediction generation) exceeds 1 hour. |
| | | **High** | Training requires a high-end server (multi-core CPU, 64GB RAM); inference time is in minutes. |
| | | **Moderate** | Training/running can be completed on a standard desktop computer or cloud virtual machine (<= 16GB RAM) in <1 hour. |
| | | **Low** | Training/running is near-instantaneous (seconds/minutes) and executable on a standard laptop CPU using basic statistical software. |

| | | | |
|---|---|---|---|
| **Data Requirements** | This dimension assesses the volume, granularity, and variety of input data needed to train the model effectively. | **Very High** | Requires multi-source, high-granularity data (e.g., electronic health records, high-resolution remote sensing, mobility data). The total number of observed case records/data points. The number of observations (Volume Metric) used for training is >250,000. |
| | | **High** | Requires high-quality, continuous, time-series data with at least 5 covariates (e.g., climate, ENSO, and social data). The number of observations used for training is between 50,001 and 250,000. |
| | | **Moderate** | Requires local, standardised surveillance data and 2-3 standard environmental covariates (e.g., rainfall, temperature). The number of observations used for training is between 10,001 and 50,000. |
| | | **Low** | Requires standard time-series data (e.g., case counts) with minimal external covariates or simple lagged variables. The number of observations used for training is less than or equal to 10,000. |

#### Appendix Table D2: Quality Assessment Questionnaire

| **Item #** | **Questions** |
|---|---|
| Q1 | The paper addresses aims and objectives. |
| Q2 | Is the setting and population clearly defined? (country) |
| Q3 | Explicitly describe the origin of the input source data, with references |
| Q4 | Is the model structure clearly described and appropriate for the research question? |
| Q5 | Is the prediction made for an observation not part of the data sample (out-of-sample validation)? |
| Q6 | Describe the model performance evaluation method used, with justification |
| Q7 | Describe the time scale/horizon |
| Q8 | Are data limitations discussed? |
| Q9 | Are the study results discussed in context, and generalisability considered? |

### Appendix Table D3: Quality Assessment

| First Author, Year | Q1 | Q2 | Q3 | Q4 | Q5 | Q6 | Q7 | Q8 | Q9 | Quality |
|---|---|---|---|---|---|---|---|---|---|---|
| Addawe, 2023 | G | G | P | G | F | F | G | P | G | M |
| Agarwala, 2024 | G | G | G | G | P | F | P | G | G | H |
| Anggraeni, 2021 | F | G | P | G | P | P | P | F | F | L |
| Anggraeni, 2024 | G | G | P | G | G | F | G | G | G | H |
| Appice, 2020 | G | G | G | G | G | F | G | F | G | VH |
| Baker, 2021 | G | G | G | G | P | G | P | P | G | M |
| Baquero, 2018 | G | G | G | F | G | G | G | P | G | H |
| Benedum, 2020 | G | G | G | F | G | G | G | G | G | VH |
| Bogado, 2021 | G | G | G | G | G | G | F | P | G | H |
| Carvajal, 2018 | G | G | F | F | G | F | G | P | G | H |
| Chakraborty, 2019 | G | G | G | G | F | G | G | P | F | H |
| Chakraborty, 2020 | F | G | F | F | G | P | P | P | F | L |
| Chumpu, 2019 | F | G | F | G | G | F | G | G | G | H |
| Dhaked, 2025 | F | G | G | G | G | F | F | P | G | H |
| Doni, 2020 | F | G | P | F | G | P | G | P | F | L |
| Guo, 2017 | G | G | F | P | G | G | G | F | G | H |
| Guo, 2018 | G | G | P | G | P | P | G | F | G | M |
| Handari, 2021 | G | G | P | G | G | G | F | G | G | H |
| Hasan, 2024 | G | G | G | G | P | G | G | G | G | VH |
| Ho, 2015 | G | G | G | F | G | P | G | P | P | M |
| Jayashree, 2015 | F | G | P | G | P | P | G | P | P | L |
| Juraphanthong, 2021 | F | G | P | G | P | F | F | G | F | M |
| Kakarla, 2023 | G | G | P | G | G | G | G | F | F | H |
| Kerdprasop, 2020 | G | G | P | F | G | P | G | P | P | L |
| Khaira, 2020 | G | G | P | G | G | P | G | P | P | M |
| Koplewitz, 2022 | G | G | P | G | G | F | G | G | G | H |
| Li, 2022 | G | G | G | G | G | G | G | F | G | VH |
| Ligue, 2022 | F | G | P | G | G | G | F | P | F | M |
| Lima, 2020 | G | G | G | F | G | F | G | F | G | H |
| Liu, 2019 | G | G | F | F | G | F | G | P | F | M |
| Mahdiana, 2017 | F | G | P | F | P | G | G | P | P | L |
| Majeed, 2023 | G | G | G | G | G | G | G | P | G | VH |
| Munarsih, 2020 | G | G | P | G | P | G | P | P | P | L |
| Mustaffa, 2024 | G | G | F | G | G | G | F | P | P | M |
| Nabilah, 2023 | F | G | P | P | G | P | F | P | P | L |
| Navarro Valencia, 2021 | G | G | G | G | G | G | G | G | G | VH |
| Nguyen, 2022 | G | G | P | F | G | G | F | G | G | H |
| Othman, 2022 | F | G | P | G | G | F | G | P | F | M |
| Panja, 2023 | G | G | G | F | G | G | G | P | F | H |
| Patil, 2021 | G | G | P | F | G | G | G | G | F | H |
| Patra, 2024 | G | G | G | F | G | G | G | P | F | H |
| Pham, 2018 | G | G | P | G | G | F | G | P | P | M |
| Phung, 2014 | G | G | P | G | G | G | G | G | G | VH |
| Polwiang, 2020 | G | G | F | F | G | P | G | G | G | H |
| Prome, 2024 | G | G | P | F | G | F | P | P | P | L |
| Puengpreeda, 2020 | P | G | F | P | G | F | G | P | F | L |
| Rajendran, 2023 | F | G | G | P | G | F | G | P | P | M |
| Rangarajan, 2019 | G | G | G | G | P | F | G | G | G | H |
| Sebastianelli, 2024 | G | G | P | F | G | P | F | P | F | L |
| Sharma, 2021 | G | G | G | F | G | P | P | P | F | M |
| Shashvat, 2019 | G | G | P | G | P | F | G | P | F | M |
| Shashvat, 2023 | G | G | P | P | G | P | G | P | P | L |
| Shi, 2015 | G | G | G | F | G | F | G | P | F | H |
| Tian, 2024 | G | G | P | P | G | P | F | G | G | M |
| Tuan, 2024 | G | G | F | G | G | G | F | G | G | VH |
| Weng, 2024 | G | G | G | P | G | P | G | G | P | M |
| Xu, 2020 | G | G | P | F | G | G | G | P | G | H |
| Zhao, 2020 | F | G | G | P | G | F | G | P | F | M |
| Zhao, 2023 | G | G | F | G | G | G | G | P | F | H |

Notes: Poor (P), Fair (F), Good (G); Low (L), Medium (M), High (H), Very High (VH)
